# Orbital Magnetic Resonance Imaging of Ocular Giant Cell Arteritis: A Systematic Review and Individual Participant Data Meta-Analysis

**DOI:** 10.1101/2022.11.11.22282193

**Authors:** Konstanze V. Guggenberger, Athanasios Pavlou, Quy Cao, Ishaan J Bhatt, Qi N. Cui, Thorsten A. Bley, Hugh D. Curtin, Julien Savatovsky, Jae W. Song

## Abstract

**Objectives:** We conducted a systematic review and individual participant data meta-analysis of publications reporting the ophthalmologic presentation, clinical exam, and orbital MRI findings in ocular giant cell arteritis.

**Methods:** PubMed and Cochrane databases were searched up to January 16, 2022. Publications reporting patient-level data on patients with ophthalmologic symptoms, imaged with orbital MRI and diagnosed with biopsy-proven giant cell arteritis were included. Demographics, clinical symptoms, exam, lab, imaging, and outcomes data were extracted. Methodological quality and completeness of reporting of case reports were assessed.

**Results:** Thirty-two studies were included comprising 51 patients (females=24; median age, 76 years). Vision loss (78%) and headache (45%) were commonly reported visual and cranial symptoms. Ophthalmologic presentation was unilateral (41%) or bilateral (59%). Fundus examination most commonly showed disc edema (64%) and pallor (49%). Average visual acuity was very poor (2.28 logMAR ± 2.18). Diagnoses included anterior (61%) and posterior (16%) ischemic optic neuropathy, central retinal artery occlusion (8%) and orbital infarction syndrome (2%). On MRI, enhancement of the optic nerve sheath (53%), intraconal fat (25%), and optic nerve/chiasm (14%) was most prevalent. Among patients with monocular visual symptoms, 38% showed pathologic enhancement in the asymptomatic eye. Six of seven cases reported imaging resolution after treatment on follow-up MRIs.

**Conclusions:** Vision loss, pallid disc edema, and optic nerve sheath enhancement are the most common clinical, fundoscopic and imaging findings reported in patients diagnosed with ocular giant cell arteritis, respectively. MRI may detect subclinical inflammation in the asymptomatic eye and may be an adjunct diagnostic tool.

**Key Points:**

- Among 32 publications comprising 51 patients with biopsy-proven giant cell arteritis, vision loss, pallid disc edema, and optic nerve sheath enhancement were the most commonly reported clinical, fundoscopic and orbital MRI findings, respectively.
- Orbital MRI detected subclinical inflammation in the asymptomatic orbit in 38% of giant cell arteritis patients presenting with monocular visual symptoms.
- MRI may have a diagnostic role in patients with suspected giant cell arteritis and presenting with acute visual symptoms.

## Introduction

Ocular manifestations among patients with giant cell arteritis (GCA), a systemic vasculitis mainly affecting medium-sized and large arteries, can lead to permanent visual impairment and irreversible blindness [1]. The reported annual incidence of GCA is approximately 19.8 per 100,000 persons aged 50 years or older [2; 3] and permanent vision loss occurs in approximately 15% to 20% of these patients [4-6]. Prompt diagnosis and treatment are imperative to prevent irreversible blindness. However, the wide spectrum of ophthalmic symptoms and nonspecific systemic symptoms as well as the possibility of occult GCA without systemic symptoms [7] can make the diagnosis challenging often requiring multispecialty care coordination and a temporal artery biopsy (TAB) [8].

Black blood magnetic resonance imaging (BB-MRI) of the scalp is increasingly being used to assess inflammation of the vessel walls of the temporal and occipital arteries to assist in the diagnosis of GCA [9]. Several studies also report orbital inflammatory findings on MRI in patients with GCA with postcontrast enhancement suggesting inflammation of the ophthalmic artery vessel walls [10], optic nerve sheath [8], intraconal fat [8; 11] and optic nerve/chiasm [8; 12]. These reports suggest MRI of the scalp and orbits could provide diagnostic information for patients with suspected GCA with ischemic optic neuropathy. However, the spectrum and prevalence of orbital MRI findings and ophthalmic symptoms in large cohorts of patients with ocular GCA have not been reported.

We performed a systematic review and individual participant data meta-analysis of the literature to determine the prevalence of orbital MR findings in the setting of ophthalmologic symptoms in patients diagnosed with biopsy-proven GCA. Given the rarity of this vasculitis and patients imaged by MRI, patient-level data from published case reports/series were extracted for analysis, a methodology used for rare diseases or when there is insufficient aggregate data for a good quality review per Cochrane guidelines [13; 14].

## Methods

### Search Strategy and Study Selection

This systematic review was conducted in accordance with the Preferred Reporting Items for Systematic Reviews and Meta-Analyses (PRISMA) guidelines and registered in Prospero (CRD42021254494). PubMed and Cochrane searches were conducted without foreign language restrictions on August 14, 2020 with an updated search performed on January 16, 2022. The search terms are provided in **Supplemental Table 1**. Manual review of the citations was performed. Publications were included if the study reported a patient diagnosed with biopsy-proven GCA, presented with visual symptoms, underwent MRI of the brain/orbits, included a description of any orbital imaging findings, and patient-level data could be extracted. Institutional review board approval or written informed consent was not required due to the literature-based nature of the study. No overlapping systematic review regarding our topic was found in the Cochrane Library.

### Data extraction

Two independent reviewers (xx, xx; for blinded review) extracted the following data: study characteristics, number of patients with biopsy proven GCA, gender, age, presenting clinical signs/symptoms, ophthalmologic exam, MRI description, biopsied tissue, treatment and outcomes data. All study design types, including letters to the editor, conference abstracts, and case reports/series were included if individual data could be extracted. Corresponding authors were contacted to gather more data. Publications were evaluated for quality using a published Case Reports Critical Appraisal Tool (**Supplemental Table 2**) and completeness of reporting using the CARE guidelines [15; 16].

### Statistical Analysis

Categorical variables are expressed in counts and percentages; continuous variables are expressed in means. Strength of agreement between two reviewers for study selection was calculated by an unweighted Cohen’s κ. Visual acuity (VA) is a measure of the ability to discriminate 2 stimuli separated in space at high contrast and is a measure of visual function. For quantitative analysis, Snellen VA was converted into the logarithm of the Minimum Angle of Resolution (logMAR), which is calculated as –log (decimal acuity), per convention in the ophthalmology literature [17; 18]. Higher values indicate poorer vision on the logMAR scale. Legal blindness is defined, in part, by VA less than or equal to 1.0 logMAR [19], while “0” on the logMAR scale indicates “no vision loss” (20/20 vision). The VA of patients with very low vision or complete blindness is typically classified using the semi-quantitative scale “counting fingers,” “hand motion,” “light perception,” and “no light perception” and were assigned values of 2, 3, 4, and 5 on the logMAR scale, respectively. The average VA was reported per patient and per symptomatic orbit. SPSS v19 (IBM, Chicago, IL) was used for statistical analysis.

### Results

#### Literature Search

Initial search yielded 157 studies, which were screened by 2 independent reviewers (xx, xx- for blinded review) with moderate agreement (κ=0.79, 95% CI 0.69-0.89, p<0.0001). Full-text screen was performed on 62 articles, from which 32 articles were qualitatively reviewed (**Figure 1**).

**Figure 1:**
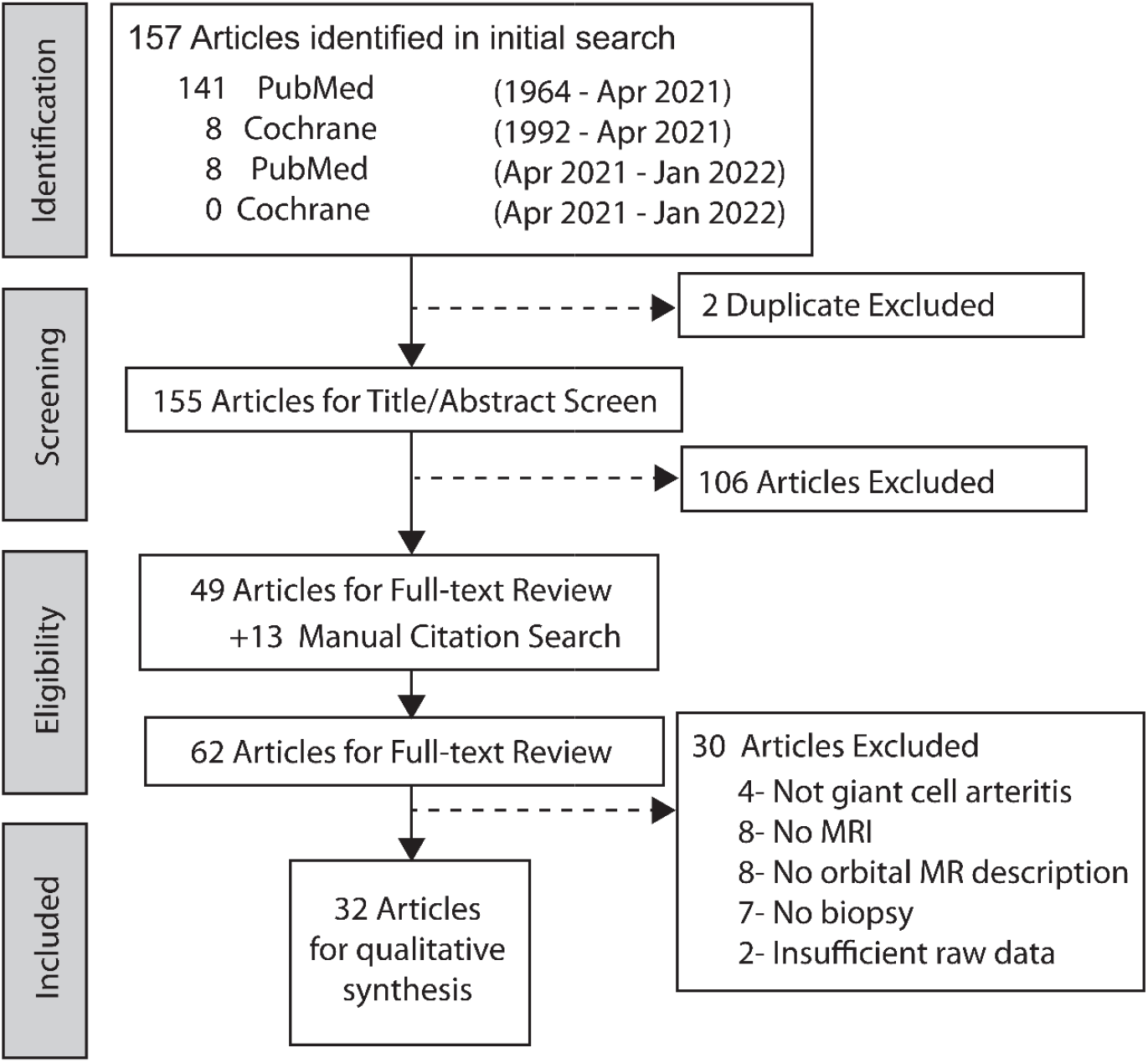
Literature Search Flowchart PubMed and Cochrane databases were searched and yielded 157 articles from which 32 met inclusion and exclusion criteria.

#### Data Synthesis

A total of 51 patients (males=27, females=24; median age=77 years (IQR 68.5, 81) [range 60-92 years]) were included in the analysis (**Supplemental Table 3**). **Figure 2A-B** shows the reported ophthalmologic symptoms/signs. Vision loss (78%, N=40/51) was the most commonly reported ophthalmologic symptom at initial presentation. Diplopia (20%, N=10/51) and orbital pain (10%, N=5/51) were also reported. On exam, an afferent pupillary defect (27%, N=14/51), reduced extraocular motility (16%, N=8/51), and proptosis (14%, N=7/51) were most commonly reported. Most common cranial symptom was headache (45%, N=23/51). When specified, headaches were temporal (64%), retro-orbital (27%), occipital (9%), or frontal (9%). Jaw claudication (20%, N=10/51), palpable temporal artery (18%, N=9/51), and scalp tenderness (18%, N=9/51) were commonly reported. Constitutional symptoms included fatigue (20%, N=10/51), weight loss (20%, N=10/51), fever (14%, N=7/51), and anorexia (10%, N=5/51) (**Figure 2C**).

**Figure 2:**
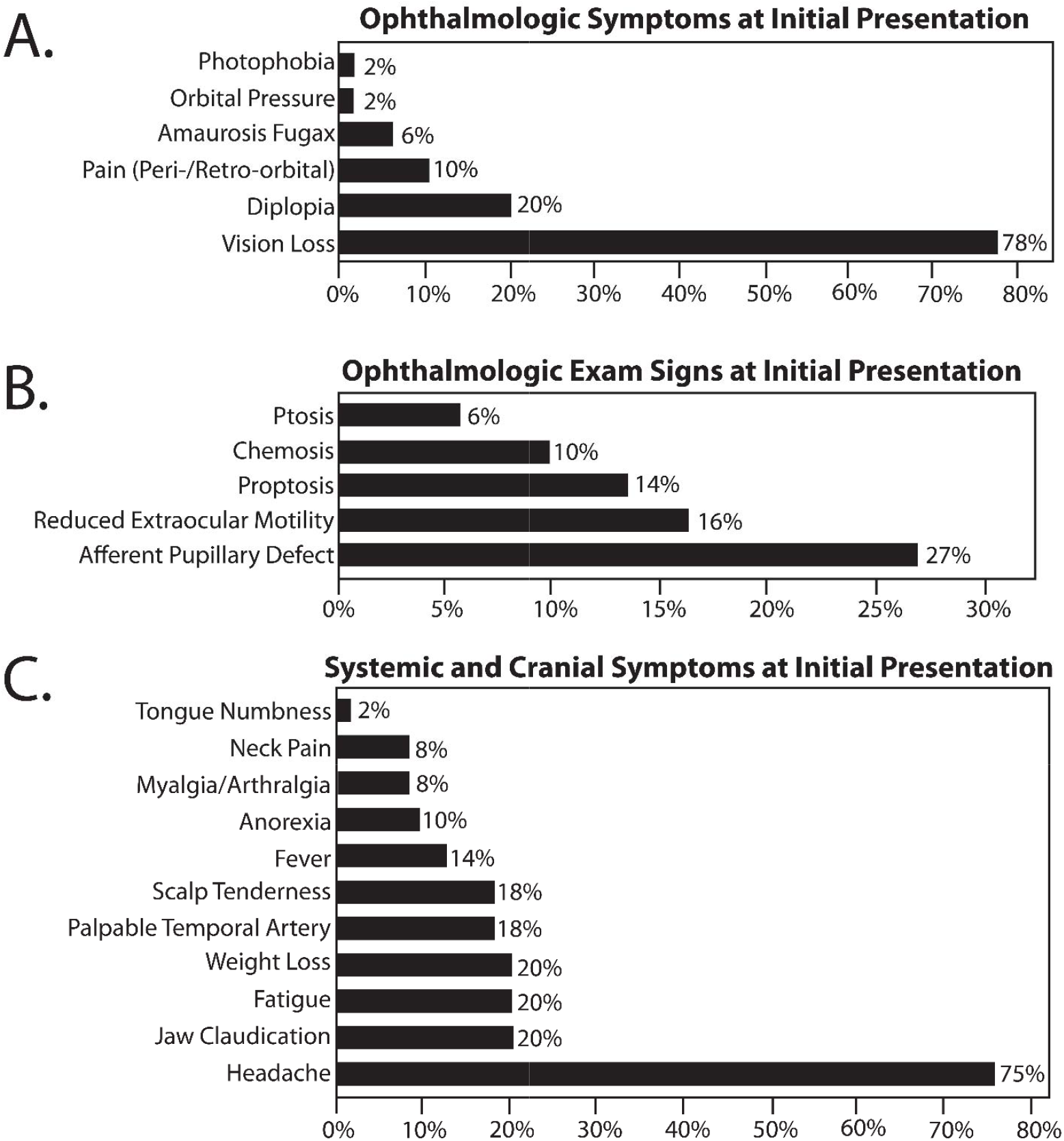
Clinical Ophthalmologic and Cranial Symptoms and Exam Signs at Initial Presentation A) Prevalence of ophthalmologic clinical symptoms B) ophthalmologic exam signs and C) cranial and systemic symptoms in patients with biopsy proven giant cell arteritis with visual symptoms.

Upon initial presentation, visual symptoms were reported to be unilateral in 41% (N=21/51) (right eye, 52%; left eye, 48%) and bilateral in 59% (N=30/51) of patients. Among patients who reported bilateral visual symptoms, 20% (N=10/51) described sequential symptom onset. On ophthalmologic exam, VA was reported for both eyes in 63% of cases (N=32/51) with an average VA of 2.28 logMAR (SD ±2.18 logMAR). Among patients who presented with unilateral symptoms with reported VA (N=11), the average VA of the symptomatic eye was 2.04 logMAR (SD ±2.10 logMAR). Among patients who presented with bilateral/sequential symptoms with reported VA (N=23), the average VA was 2.91 logMAR (SD ±2.12 logMAR).

Fundus examinations were reported in 76% of cases (N=39/51). Disc findings included disc edema (64%, N=25/39), disc pallor (49%, N=19/39), disc hemorrhage (11%, N=4/39), and disc atrophy (5%, N=2/39). Other findings included cotton wool spots (10%, N=4/39), cherry red spot (5%, N=2/39), and vascular narrowing (5%, N=2/39). Reported diagnoses included anterior ischemic optic neuropathy (61%, N=31/51), posterior ischemic optic neuropathy (16%, N=8/51), and central retinal artery occlusion (8%, N=4/51). Orbital infarction syndrome was reported in 1 case as a result of absent filling of the right ophthalmic artery [20].

The erythrocyte sedimentation rate (ESR) and c-reactive protein (CRP) were elevated in 83% (N=33/40; normal considered <20 mm/hr) and 42% (N=11/26; normal considered <0.5mg/L) of reported cases, respectively. The ESR and CRP were both normal in 8% of cases (N=2/26) [21; 22]. Five cases reported platelet counts, with thrombocytosis (above 450 × 10^9^/L) in 3 of the cases [23-25].

#### Orbital MR Imaging Findings

Neuroimaging included combined brain and orbit (76%, N=39/51) or only orbital MRIs (24%, N=12/51). Two studies used dedicated black blood MR imaging (BB-MRI) [22; 26]. Eleven cases reported the steroid-to-imaging time interval, which ranged between 1 day [22] and 12 months [11]. **Figure 3** shows the prevalence of reported pathologic MR signal detected in the orbit among patients diagnosed with ocular GCA (N=51) and a subgroup analysis among those who presented with vision loss (N=40).

**Figure 3:**
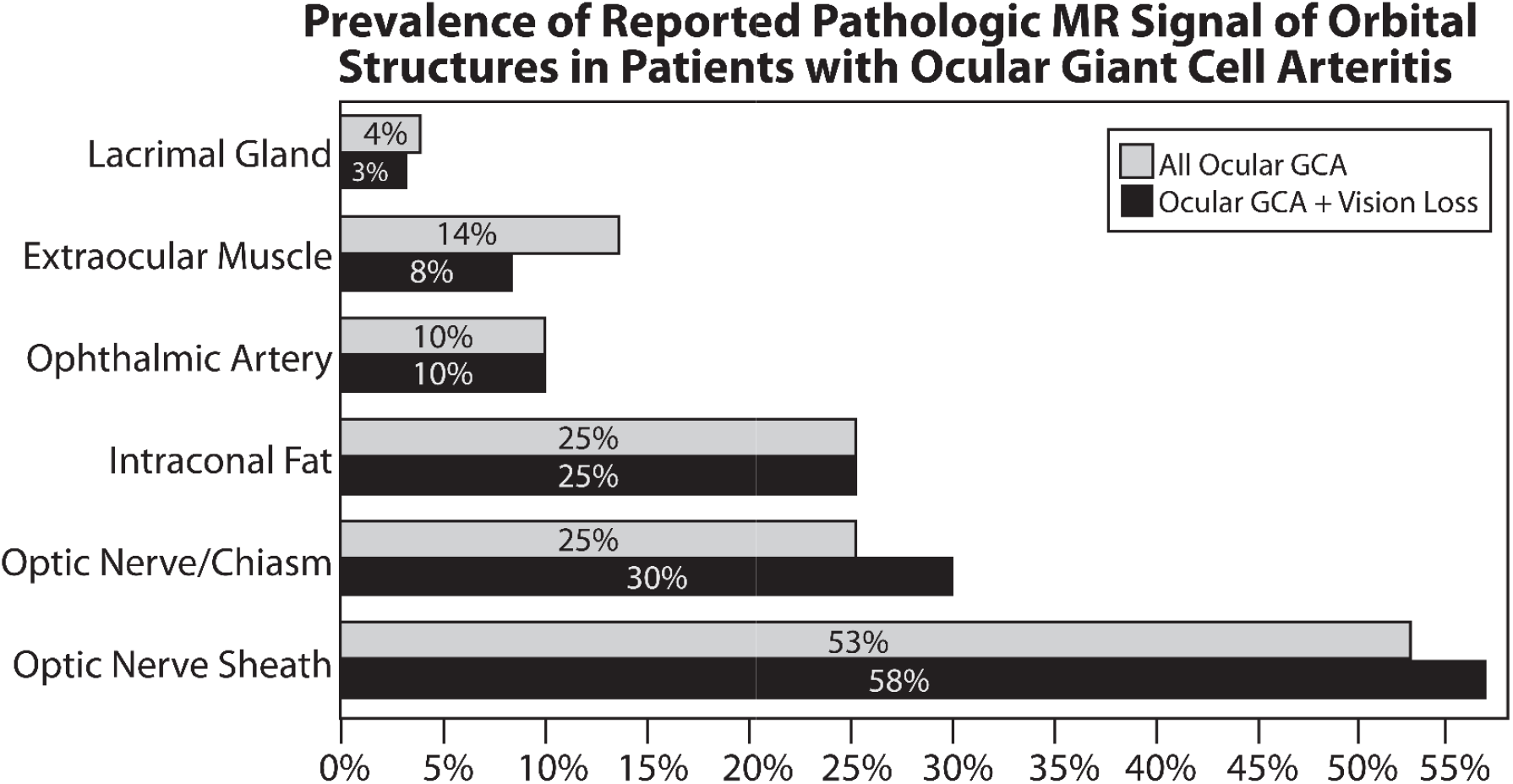
Prevalence of Reported Pathologic MR Signal of Orbital Structures in Patients with Ocular Giant Cell Arteritis Prevalence of pathologic MR signal (e.g., postcontrast enhancement, T2-weighted/STIR hyperintensity, or diffusion restriction) of orbital structures on MRI in patients with biopsy proven giant cell arteritis among all ocular GCA patients and subgroup presenting with vision loss are shown. Abbreviations: GCA, giant cell arteritis; STIR, short tau inversion recovery; MRI, magnetic resonance imaging

Pathologic enhancement on postcontrast T1-weighted imaging was the most commonly reported description with enhancement of the optic nerve sheath (53%, N=27/51), intraconal fat (25%, N=13/51), optic nerve/chiasm (14%, N=7/51), extraocular muscle (14%, N=7/51) and lacrimal gland (2%, N=1/51). When assessed, ophthalmic artery vessel wall enhancement was described in 10% (N=5/51) of cases. Diffusion restriction (8%, N=4/51)[20; 27-29] or T2/STIR hyperintensity (4%, N=2/51) [28; 30] of the optic nerve, diffusion restriction of the lacrimal gland (2%, N=1/51) [31], and enlargement of the extraocular muscle (2%, N=1/51) [21] were also reported. The optic nerve/chiasm showed any pathologic MR signal abnormality (e.g., postcontrast enhancement, T2-weighted/Short Tau Inversion Recovery (STIR), or diffusion restriction) in 25% (N=13/51) of cases of ocular GCA. Subgroup analyses with vision loss showed 30% (N=12/40) of cases revealed pathologic MR signal in the optic nerve/chiasm. **Figure 4A-F** illustrates representative MRI findings on a dedicated orbital MRI and BB-MRI from patients diagnosed with ocular GCA from the authors’ institutions.

**Figure 4:**
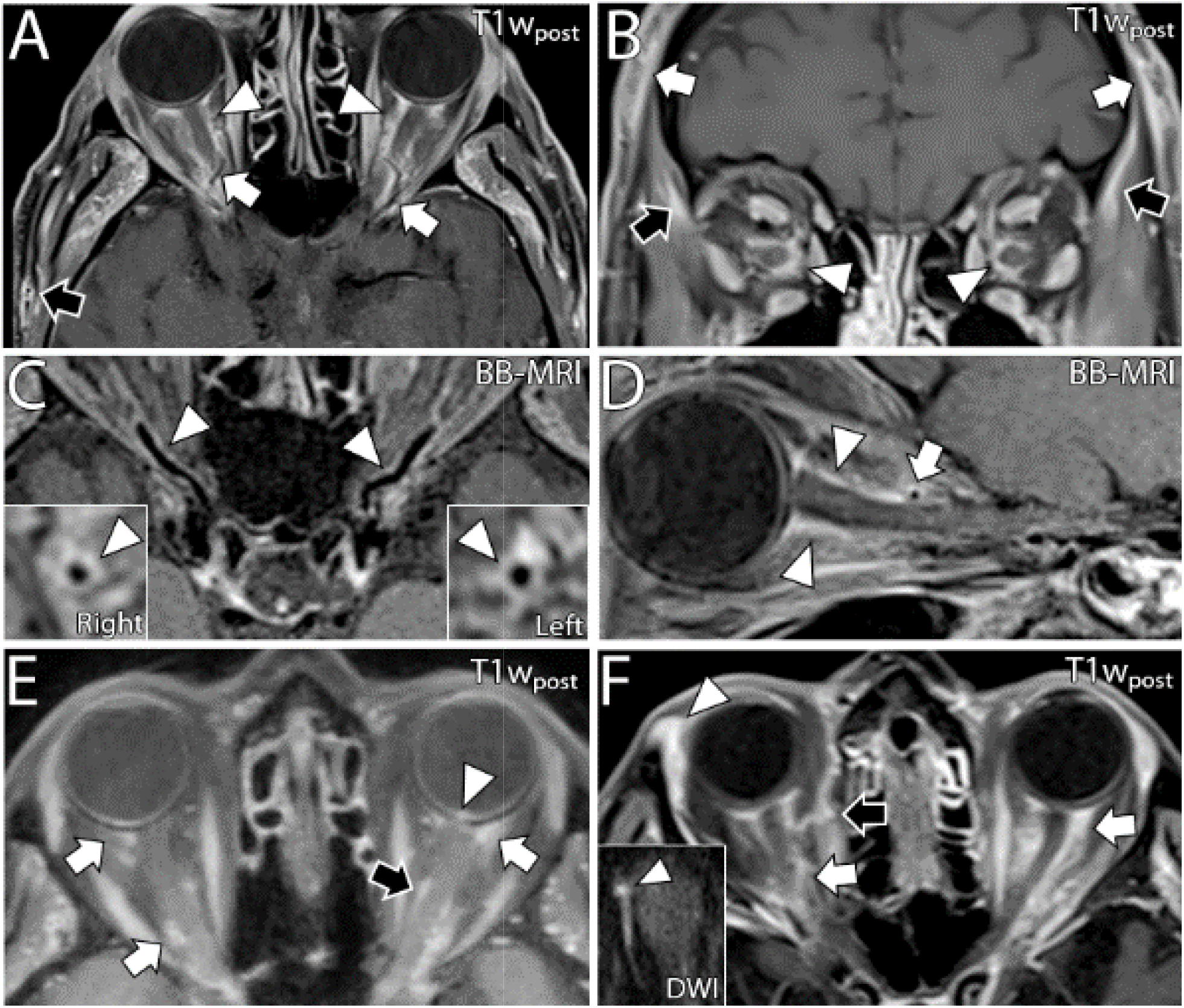
Example Orbital and Black-Blood MR Images of Patients with Ocular Giant Cell Arteritis (A) Axial T1-weighted postcontrast fat suppressed image shows avid enhancement of the intraorbital fat (arrowheads) and ophthalmic arteries (white arrows). The right superficial temporal artery also showed vessel wall enhancement (black arrow). (B) Coronal T1-weighted postcontrast fat suppressed image of the same patient shows enhancement of the optic nerve sheath (arrowheads). Enhancement of the temporalis muscle (black arrows) and subcutaneous fat of the scalp associated with the frontal branches of the superficial temporal arteries (white arrows) was also present. (C) Axial high spatial resolution BB-MRI imaging revealed vessel wall enhancement of the ophthalmic arteries (arrowheads). Insets show orthogonal images of the thickened and enhancement ophthalmic artery vessel walls (arrowheads), bilaterally. (D) Sagittal-oblique high spatial resolution BB-MRI of the right optic nerve shows enhancement of the optic nerve sheath (arrowheads) and ophthalmic artery vessel walls (arrow). (E) Axial T1-weighted postcontrast fat suppressed image shows enhancement of the left optic nerve head (arrowhead), left optic nerve sheath (black arrow), and bilateral intraconal fat (white arrows). (F) Axial T1-weighted postcontrast fat suppressed image shows enhancement and asymmetric enlargement of the right lacrimal gland (arrowhead), which also showed focal diffusion restriction (inset, arrowhead). Enhancement of the bilateral intraconal fat (white arrows) were present as well as linear enhancement that tracked along the right anterior ethmoidal artery (black arrow). Abbreviations: BB-MRI, black-blood MRI; T1w, T1-weighted postcontrast; DWI, diffusion weighted imaging

#### Concordance of Monocular Visual Symptoms and Orbital MRI Findings in the Ipsilateral Symptomatic versus Contralateral Asymptomatic Orbit

Subgroup analysis of patients presenting with monocular visual symptoms (N=21) showed pathologic MR signal of orbital structures in the symptomatic eye in 91% (N=19/21) of cases. The most commonly involved orbital structure was the optic nerve sheath (38%, N=8/21) [23; 29; 32-35] followed by the optic nerve/chiasm [24%, N=5/21; N=2 showed diffusion restriction [20; 29], N=1 showed STIR hyperintensity [30], and N=2 showed enhancement [8; 35]], intraconal fat (24%, N=5/21), extraocular muscles (19%, N=4/21), ophthalmic artery (19%, N=4/21), and lacrimal gland (10%, N=2/21).

Pathologic orbital MRI findings in the asymptomatic orbit in patients with monocular visual symptoms were also reported in 38% (N=8/21) of cases (**Figure 5**). The most common pathologically enhancing orbital structures on MRI in the asymptomatic orbit among patients with monocular vision loss were the optic nerve sheath (24%, N=5/21), intraconal fat (14%, N=3/21), and ophthalmic artery (14%, N=3/21).

**Figure 5:** Orbital MRI findings in Patients with Ocular Giant Cell Arteritis and Monocular Visual Symptoms Prevalence of pathologic MR signal of orbital structures in the symptomatic versus asymptomatic eye is shown.

#### Pathologic Orbital MR Enhancement in GCA patients Without Visual Symptoms

Two studies included cases that showed orbital MR findings in patients without visual symptoms/signs. Donaldson et al reported 3 TAB proven cases of GCA in patients without visual symptoms but with bilateral ophthalmic artery vessel wall enhancement [34]. These patients reportedly presented with cranial symptoms of GCA (e.g., headaches, scalp tenderness, jaw claudication).

Geiger et al reported 10 TAB proven cases of GCA in patients without visual symptoms, among which 4 showed bilateral ophthalmic artery enhancement and 2 showed unilateral ophthalmic artery enhancement; the remaining 4 cases showed no ophthalmic artery enhancement [26].

#### Diagnosis and Outcomes

The reported time-interval from presentation until diagnosis ranged from 2 weeks [23] to 3 months [21]. A diagnosis of GCA was confirmed by TAB (90%, N=46/51), extraocular muscle (2%) [21], or optic nerve sheath (2%) [36] biopsies. One study reported “biopsy-proven” but did not specify what tissue was biopsied [32]. In one case, bilateral TABs were reported to be negative but positive in the extraocular muscles [21].

Clinical outcomes were reported in 48% of the cases. Follow-up ranged from 7 days [35] to 3 months [37]. When reported, ophthalmologic outcomes improved (9%), remained stable (33%), or worsened (7%) at follow-up. Seven cases reported follow-up MR exams, among which 6 cases [21; 25; 30; 35; 37; 38] reported resolution and 1 case [39] reported no improvement.

In 24% (N=12/51) of cases, treatment was delayed due to misclassification. Alternative diagnoses included sinusitis, lymphoma, optic neuritis, non-arteritic ischemic optic neuropathy/transient ischemic attack/embolism, infectious conjunctivitis, episcleritis, diabetic neuropathy, and infectious orbital cellulitis.

#### Quality and Completeness of Reporting

A critical appraisal checklist for case reports and the CARE (Case Report) Guideline was used to assess the quality and the reporting completeness of case reports (**Supplemental Figure 1A-B**), respectively [15; 16]. Reporting the “clinical findings and timeline” was most complete (100%) and “patient perspective” was not fulfilled in any publication. “Informed consent” was obtained in 19% of publications.

## Discussion

We summarize patient-level data from case reports/series of patients diagnosed with biopsy-proven giant cell arteritis, presenting with visual symptoms, and imaged by orbital MRI. Vision loss was the most common presenting ophthalmologic symptom. On orbital MRI, enhancement of the optic nerve sheath, intraconal fat and optic nerve/chiasm were the most common pathologic imaging findings. When assessed, imaging also revealed enhancement of the ophthalmic artery vessel walls in 10% of cases. Moreover, among 38% of patients with monocular visual symptoms, the contralateral asymptomatic eye also revealed pathologic enhancement. These results add to the literature by informing clinicians about the range of orbital imaging manifestations for this vasculitis as well as suggesting a potential diagnostic role for MR imaging in patients with ocular giant cell arteritis.

Thrombosis of the posterior ciliary arteries (PCA) is a hallmark of GCA [40]. Vision loss occurs secondary to ischemic infarction of the optic nerve or the retina due to thrombosis by granulomatous inflammation of the PCA or the central retinal artery, respectively. In a prospective cohort of 66 biopsy proven GCA patients with vision loss, fluorescein angiography revealed occlusion of the PCAs or moderate to marked delay in perfusion of the choroidal vascular bed [1]. The frequent involvement of the medial and lateral long PCAs, which course parallel to the optic nerve sheath, could explain optic nerve sheath and intraconal fat enhancement on imaging. Reports of isolated optic nerve sheath enhancement raise the possibility of inflamed vessels within the perioptic subarachnoid space, which may be below the spatial resolution of MR imaging. Scanning electron microscopy studies on autopsy specimens report arachnoid trabeculae and septae that divide this perioptic subarachnoid space through which lymphatic and blood vessels traverse [41]. Optic nerve enhancement may be explained by involvement of the central retinal or cilioretinal artery. Finally, inflamed branches of the internal carotid artery (ophthalmic segment) or proximal ophthalmic artery could explain chiasmal enhancement. In fact, studies report variable intracranial internal carotid artery vessel wall enhancement in patients with GCA [22]. Attention to the anatomy of orbital pathologic enhancement may yield information about orbital vascular territories and aid with risk-stratification.

Interestingly, 38% of cases with monocular visual symptoms also revealed enhancement of the optic nerve sheath (24%), intraconal fat (14%), and ophthalmic artery (14%) in the asymptomatic orbit. This result suggests imaging may detect subclinical ischemia. Hayreh et al reported that among 27 of 85 prospectively recruited GCA patients presenting with simultaneous vision loss, fundus exams revealed evidence of older changes in the contralateral eye as well as the newly involved eye [42]. Patients may not become aware of partial unilateral vision loss until the second eye is also affected, which may result in a presentation of simultaneous rather than sequential vision loss [42]. The incidence of sequential versus simultaneous vision loss likely depends on when the GCA diagnosis is made and how soon the patient is evaluated. In our review, one case reported unilateral vision loss and optic disc blurring only in the symptomatic eye despite bilateral optic nerve sheath enhancement on MRI [33]. A negative fundus exam in the setting of positive MRI findings in the asymptomatic eye raises the possibility of early ischemic changes and possibly developing posterior ischemic optic neuropathy, a less common etiology of vision loss in GCA. Given that a primary goal for treating ocular GCA is preserving vision in the asymptomatic eye, MRI may have an adjunct diagnostic role to the fundus exam.

The role of orbital MRI for evaluating suspected cranial GCA is an area of future investigation. The European League Against Rheumatism recommends the use of imaging for diagnosing and monitoring GCA. BB-MRI can evaluate the scalp artery vessel walls if ultrasound is not available or inconclusive [43]. In the presence of visual symptoms, a comprehensive MRI assessment of both orbits in addition to BB-MRI for scalp artery imaging could be valuable in the acute setting. However, in the absence of visual symptoms, the role of orbital MRI as a screening tool for diagnosis is unclear and may not be cost-effective. Additional areas of future investigation include whether MRI could help determine when corticosteroids/immunomodulatory treatments could be tapered, if MRI can detect relapse or whether MRI has a prognostic role for determining ophthalmologic outcomes. Several cases [21; 30; 35; 37; 38] reported resolution of pathologic enhancement on orbital MRI and improvement in visual function/symptoms while another case [25] reported no visual functional improvement despite imaging resolution. Alternatively, optical coherence tomography/-angiography can image the retinal layer and assess microvascular changes, respectively. These tools may be more precise in determining ocular GCA outcomes and visual potential by evaluating the association between capillary flow density and retinal nerve fiber layer thickness over time [44; 45]. These questions are areas of future prospective investigations.

There are limitations to this study. First, a selection bias in the cases that underwent brain/orbital MR imaging likely resulted in an overestimation in the prevalence of pathologic orbital findings. Orbital MRIs are not commonly performed in patients with suspected cranial GCA, especially in the absence of ophthalmologic symptoms. Moreover, few institutions may have access to MRIs, particularly in the emergency department. Nevertheless, a summary of the reported cases provides new knowledge about the spectrum of orbital imaging findings in patients with ocular GCA and adds to the literature about the potential role of MRI in patients with ocular GCA. Second, TAB is an imperfect gold standard for the diagnosis of GCA [46]. As biopsy proven GCA was an inclusion criterion for this review, the true prevalence of pathologic orbital imaging findings in ocular GCA may not be accurate. Third, patients diagnosed with GCA but without visual symptoms may not have been imaged with an orbital MRI; this subgroup was not an inclusion criterion of this study. Hence, the true prevalence of pathologic orbital findings in cranial GCA, with or without ophthalmic symptoms, is not established in this review. Fourth, the ophthalmologic work-up and reporting of ocular examinations varied, limiting the completeness of data extraction. Attempts to contact authors were made with variable success. Fifth, the MRI protocol was not detailed in most reports. Variations in pulse sequences and image interpretation may limit accuracy. Moreover, most studies did not use high spatial resolution BB-MRI and likely were not able to evaluate the ophthalmic artery vessel walls. A purpose of this study therefore is to highlight the importance of evaluating orbital anatomy in suspected GCA cases with ophthalmologic symptoms.

In conclusion, we present individual-participant level data on orbital MRI findings in patients with biopsy proven ocular giant cell arteritis. Vision loss is a severely debilitating complication of ocular giant cell arteritis, which can be minimized or prevented if recognized early and treated promptly. Understanding the spectrum of orbital MRI findings in these cases is important for the radiologist as scalp black-blood MRI, which can include the orbital anatomy, is becoming increasingly adopted clinically in the diagnostic evaluation of these patients.

## Data Availability

All data produced in the present study are available upon reasonable request to the authors. This study has been submitted to the journal European Radiology.

## Abbreviations

GCA: giant cell arteritis
BB-MRI: black-blood MRI
TAB: temporal artery biopsy
VA: visual acuity

## Acknowledgements

This work is supported by the American Heart Association Career Development Award 938082 (JWS) and NIH/NEI K08EY029765 (QNC). Dr. Jae W. Song is the guarantor of this study.

