## Supplement for "Orbital Magnetic Resonance Imaging of Ocular Giant Cell Arteritis: A Systematic Review and Individual Participant Data Meta-Analysis"

**Supplemental Figure and Table:**

**Supplemental Figure 1:** Case Report Critical Appraisal and Completeness of Reporting based on the CARE Case Report Guidelines

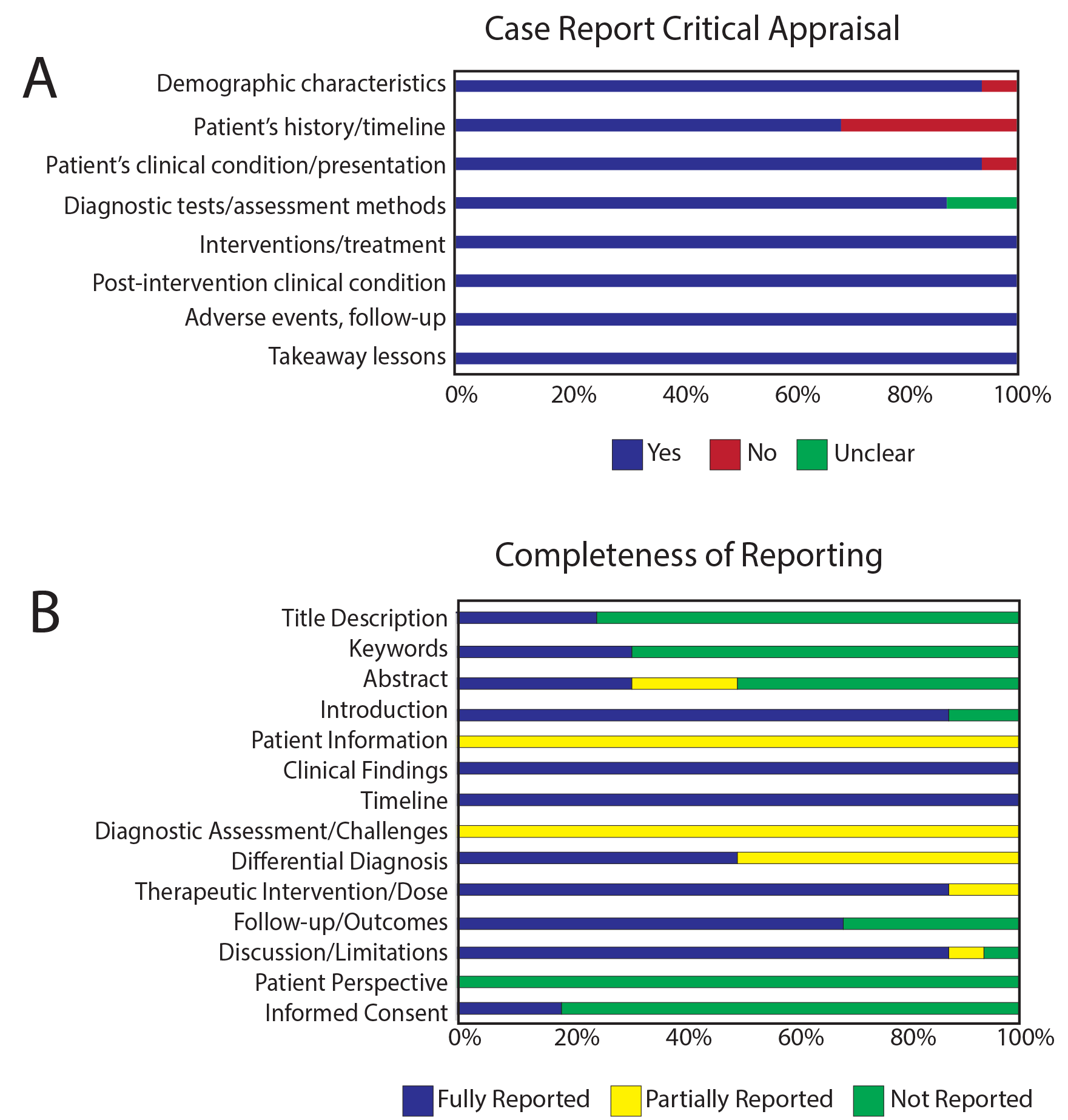

**Supplemental Table 1:** Search Strategy in PUBMED

("Giant Cell Arteritis"[Mesh] OR Giant Cell Arteritis OR Giant Cell Arteritides OR Horton OR Horton's OR Hortons OR Temporal Arteritis OR Temporal Arteritides”) AND ("Orbital Diseases"[Mesh] OR Orbital OR Retrobulbar OR Intraconal OR “Oculomotor Muscles"[Mesh] OR Oculomotor Muscle* OR Extraocular Muscle* OR Musculus Orbitalis OR Rectus Muscle OR Superior Oblique OR Inferior Oblique OR "Orbit"[Mesh] OR Orbit* OR Eye socket OR "Optic Nerve"[Mesh] OR Optic nerve* OR Cranial Nerve* II OR Second Cranial Nerve* OR "Ophthalmic Artery"[Mesh] OR Ophthalmic arter* OR "Ciliary Arteries"[Mesh] OR Ciliary arter*­ OR "Lacrimal Apparatus"[Mesh] OR Lacrimal) AND ("Magnetic Resonance Imaging"[Mesh] OR Magnetic OR NMR OR MR OR MRA OR MRI OR Zeugmatography OR Chemical shift OR Proton spin OR Spin echo)

**Supplemental Table 2:** Case Reports Critical Appraisal Tool [16]

1. Were patient’s demographic characteristics clearly described?
2. Was the patient’s history clearly described and presented as a timeline?
3. Was the current clinical condition of the patient on presentation clearly described?
4. Were diagnostic tests or methods and results clearly described?
5. Was the intervention(s) or treatment procedure(s) clearly described?
6. Was the post-intervention clinical condition clearly described?
7. Were adverse events (harms) or unanticipated events identified and described?
8. Does the case report provide takeaway lessons?

Supplemental Table 3: Individual Patient-level Data with Clinical Presentation, Fundus Exam, and MRI Findings

| Author/ Year | Sex | Age | Vision Loss (Y/N), Side (L/R/BL) | Headache (Y,N) | R, VA | L, VA | R, fundus exam | L, fundus exam | R, orbital MRI^a^ | L, orbital MRI^a^ |
| --- | --- | --- | --- | --- | --- | --- | --- | --- | --- | --- |
| Nassani et al 1995 [38] | F | 69 | N | Y | NR | NR | NR | NR | ONS, ICF | ONS, ICF |
| Lee et al 1999 [12] | F | 82 | Y, BL | N | 20/400 | NLP | DP, DE | DP, DE | ONS | ONS |
|  | F | 86 | Y, BL | N | CF | NLP | DP | DP, DE, H | ON | ON |
|  | M | 80 | Y, R | Y | CF | 20/20 | Normal | Att | ON | ON |
| Joelson et al 2000 [37] | M | 69 | N | Y | NR | NR | NR | NR | EOM | Normal |
| Lee et al 2001 [21] | M | 70 | Y, BL | Y | 20/200 🡪 NLP | 20/40 🡪 20/100 | OAT | DE | ICF, EOM^b^ | ICF, EOM^b^ |
|  | M | 69 | N | Y | 20/20 | 20/20 | NR | NR | ONS | ONS, EOM |
|  | F | 72 | N | N | 20/30 | 20/25 | Att | Att | EOM^c^ | Normal |
|  | M | 82 | N | Y | 20/60 | 20/70 | Normal | Normal | EOM^d^ | Normal |
| López et al 2001 [47] | M | 73 | Y, R | Y | CF | 0/20 (cataract) | DP, DE | NR | ON | Normal |
| Garcia-Porrua et al 2003 [32] | M | 65 | Y, L | Y | NR | NR | Normal | Normal | Normal | ONS, ICF |
| Morgenstern et al 2003 [36] | M | 83 | Y, BL | N | NLP | 20/50 | DP, DE | DP, DE | ONS | ONS |
| Botella-Ortiz et al 2004 [48] | F | 81 | Y, BL | Y | NR | NR | Normal | Normal | EOM | EOM |
| Garcia-Porrua et al 2005 [30] | F | 82 | Y, R | Y | HM | NR | DP, DE, H | NR | ON^e^ | Normal |
| Biotti et al 2011 [29] | F | 80 | Y, R | N | NR | NR | DP, DE, CRS | Normal | ON^f^ | Normal |
| Liu et al 2013 [23] | F | 83 | Y, BL | Y | HM 🡪 NLP | 20/30 🡪 NLP | Normal | Normal | ONS | ONS |
|  | F | 68 | Y, L | N | 20/20 | 20/25 🡪 NLP | Normal | DP, DE, H | Normal | ONS |
| Hittinger et al 2014 [35] | M | 78 | Y, L | Y | NR | 20/200 | NR | NR | Normal | ONS, ICF, EOM, ONC, ON, LG, OA |
| Mitchell et al 2014 [49] | M | 68 | N | Y | Normal | Normal | CWS | Normal | ICF | ICF |
| Attaseth et al 2015 [50] | M | 78 | Y, BL | Y | LP | HM | DP, DE | DP, DE | ONS, ICF | ONS, ICF |
|  | M | 70 | Y, R | Y | HM | Normal | DP, DE | NR | Normal | Normal |
|  | M | 75 | Y, BL | Y | NLP | NLP | DP, DE | DP, DE | ONS, ICF | ONS, ICF |
|  | F | 63 | Y, R | Y | CF | NR | DP, DE | NR | Normal | ONS^g^ |
|  | M | 68 | Y, L | Y | NR | Normal | DP, DE | NR | Normal | Normal |
| Chen et al 2015 [51] | F | 79 | Y, BL | N | HM 🡪 LP | 20/40 🡪 LP | DP, DE | DP, DE | ONS | ONS |
| Kornberg et al 2015 [33] | M | 67 | Y, L | N | 20/30 | 20/100 | Normal | DE | ONS, ICF, ONC | ONS, ICF, ONC |
| D’Souza et al 2016 [8] | M | 60 | Y, L | Y | 20/40 (cataract) | CF | NR | DP, DE | Normal | ON |
|  | M | 83 | Y, BL | N | NLP | CF | NR | DP | Normal | ONS |
|  | M | 63 | Y, BL | N | LP | LP | DE | DE | ONS, ONC | ONS, ONC |
| Kim et al 2016 [20] | M | 76 | N | Y | NR | NR | NR | NR | ON^f^ | Normal |
| Ross et al 2017 [27] | M | 73 | Y, BL | Y | NLP | 20/50 | NR | Normal | ONC^h^ | Normal |
| Albarrak et al 2018 [28] | M | 80 | Y, BL | Y | NLP | NLP | Normal | Normal | ON^f, j^, OA^i^ | OA^i^ |
| AlShaker et al 2018 [11] | F | 80 | Y, BL | Y | CF 🡪NLP | 20/150 | DP, DE | DE | ONS, ICF | ONS. ICF |
| Heraud et al 2018 [31] | M | 83 | N | N | NR | NR | NR | NR | LG^f^ | Normal |
| Choi et al 2019 [39] | F | 81 | Y, BL | Y | NLP | NLP | DP, DE, CWS | DP, DE | ONS | ONS |
|  | F | 68 | Y, BL | Y | 20/100 | 20/80 🡪 20/100 | DP, DE, CWS | DP, DE, CWS | Normal | Normal |
| Donaldson et al 2020 [34] | F | 79 | Y, R | N | NR | NR | DP, DE | NR | OA | OA |
|  | M | 92 | Y, L | N | NR | NR | NR | NR | OA | OA |
|  | M | 73 | N | Y | NR | NR | NR | NR | OA | OA |
| Mears et al 2020 [52] | M | 84 | Y, BL | N | NLP | 20/60 | DE, H | DE, H | Normal | ONS |
| Serrano Alcalá et al 2020 [53] | F | 62 | Y, BL | Y | NLP | NLP | DE | DE | ONS, ICF | ONS, ICF |
| Emami et al 2021 [24] | M | 60 | N | Y | 20/20 | 20/20 | DE, CWS | NR | ONS, ICF | ONS, ICF |
| Gospe et al 2021 [22] | F | 79 | Y, BL | N | NLP | NLP | NR | NR | ONS | ONS |
|  | F | 83 | Y, R | Y | NR | NR | Normal | Normal | Normal | Normal |
|  | F | 87 | Y, BL | Y | NR | NR | Normal | Normal | Normal | Normal |
|  | M | 75 | N | N | NR | NR | NR | NR | ONS | ONS |
|  | F | 64 | Y, L | N | NR | NLP | NR | NR | ONS | ONS |
|  | F | 67 | Y, BL | N | NR | NR | NR | NR | ONS | ONS |
|  | F | 76 | Y, L | N | NR | NR | NR | NR | ONS | ONS |
| Panneerselvam et al 2021 [54] | F | 73 | Y, L | N | 20/20 | NLP | NR | CRS | ONS, ICF | ONS, ICF |
| Tashiro et al 2021 [25] | F | 76 | Y, BL | Y | Normal | 20/25 | Normal | DE | ONS, ICF | ONS, ICF, ON |

^a^All MR signal changes are postcontrast enhancement unless otherwise indicated in the footnotes.

^b^Reported as poorly defined abnormal signal intensity in the orbits most prominent in the medial recti of both eyes

^c^Right medial rectus muscle showed “signal abnormality” with possible contrast enhancement

^d^Enlargement of the extraocular muscles of the right orbit

^e^STIR hyperintensity

^f^Diffusion restriction

^g^Confirmed via personal author communication.

^h^Reported diffusion restriction and FLAIR hyperintensity of the right optic nerve

^i^Ophthalmic artery not visualized on magnetic resonance angiography

^j^T2 hyperintensity

Abbreviation: L, left; R, right; N, no; BL, bilateral; M, male; F, female; VA, visual acuity; ONC, optic nerve/chiasm; ON, optic nerve; OA, ophthalmic artery; ONS, optic nerve sheath; ICF, intraconal fat; LG, lacrimal gland; EOM, extraocular muscle; CF, counting fingers; HM, hand motion; LP, light perception; NLP, no light perception; DE, disc edema; DP, disc pallor; H, hemorrhage; CRS, cherry red spot; CWS, cotton wool spots; Att, attenuated/narrowed arterioles; OAT, optic atrophy; NR, not reported­

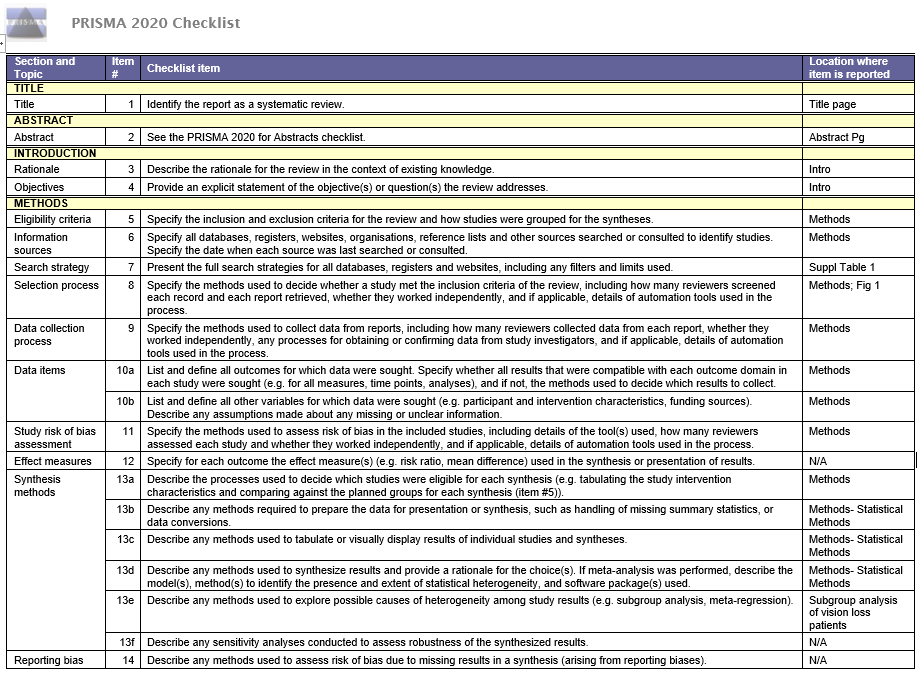

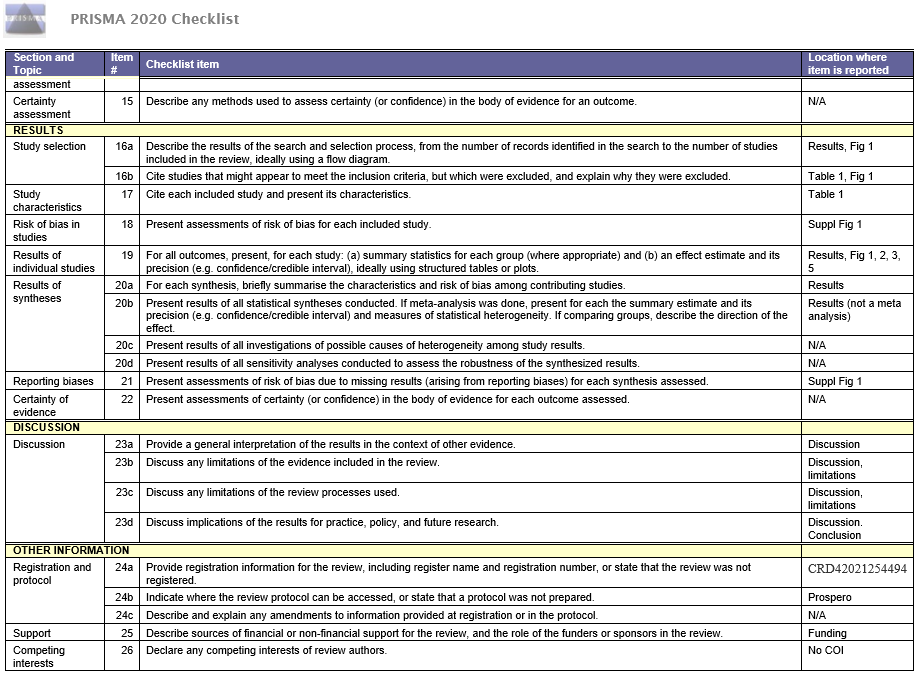

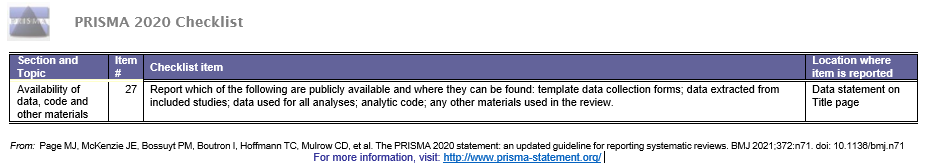
